# Typhoon Eye Effect vs. Ripple Effect: The Role of Family Size on the Mental Health under the COVID-19 Pandemic in Pakistan

**DOI:** 10.1101/2020.11.19.20229856

**Authors:** Tooba Lateef, Jiyao Chen, Muhammad Tahir, Teba Abdul Lateef, Bryan Chen, Jizhen Li, Stephen X. Zhang

## Abstract

**Background:** The recent outbreak of COVID-19 impacts the mental health of people worldwide. The mental conditions and the associated predictors of adults in Pakistan, the fifth most populous country in the world, during the COVID-19 remains understudied. We aim to investigate distress, anxiety and overall mental health and their associated predictors among Pakistani adults in this pandemic. We specifically examine the mental health issues based on the distance to the epicenter, a predictor that has revealed opposing evidence in other countries based on the theories of typhoon eye effect and ripple effect. The samples consist of 601 adults who were surveyed online about 2.5 months into the outbreak across Pakistan with varying distance to the epicenter of COVID-19 of Karachi in Pakistan.

**Results:** The results showed that 9.2% and 19.0% of the participants surpassed the cut-off of distress and anxiety disorders, respectively. Overall, the distance to the epicenter positively predicted the mental health of adults in Pakistan, and family size negatively moderated this effect. The distance to the epicenter negatively predicted distress and anxiety disorders for adults in large families, which are quite common in Pakistan.

**Conclusion:** The evidence of the study interestingly finds the prediction of the mental health of people by their distance to the epicenter depends on the family. The evidence of this study can help to provide the initial indicator for mental health care providers to screen vulnerable groups in Pakistan, a populous country that continues to struggle to cope with the COVID-19 pandemic.

## INTRODUCTION

In Pakistan, the first case of COVID-19 appeared on February 26, 2020 in Karachi, the largest city and the financial, industrial, and trading hub of the country. The initial cases were imported to Karachi from abroad but later on community spread started, and Karachi became the initial epicenter of the virus infection in Pakistan [1]. As COVID-19 spreads, a psychological panic among the public happened across the country, as it has happened in other countries as well such as China, Iran, Italy, Peru, Bolivia, etc. [2-8]. For instance, one study of students has shown moderate anxiety and distress as pandemic affects daily life activities in Pakistan [9].

While it seems that people closer to the center of any disastrous event would be affected more and in turn might have more mental issues [10] and the effect of catastrophic event declines for people with greater geographical distance to the epicenter, known as the “ripple effect” [11]. However, findings have demonstrated an opposite effect referred to as “psychological typhoon eye effect”. In the 2008 Wenchuan earthquake, it was first observed that people closer to the area of crisis felt calmer [12]. Later on the same phenomenon was observed during different public health emergencies [13-15], and also in the epidemic of Severe Acute Respiratory Syndrome (SARS) [10].

In the ongoing COVID-19 pandemic, the opposite theories of psychological typhoon eye effect and ripple effect have been reported [3, 16-19]. The survey findings of China and Brazil documented that people far from the epicenter had worse anxiety and distress [18-20]. Though a survey in China on healthcare staff including nurses indicated ripple effect [21]. Few other studies also reported psychological disturbances in several countries but the association with geographical distance was not assessed and hence limited the explanation of both theories [5, 22]. Up till now there has been no research conducted in the general population of Pakistan to assess anxiety and distress.

Therefore, the present study aims to study the impact of COVID-19 on the two opposite theories of typhoon eye effect and ripple effect in Pakistan. Moreover, this study is the first to examine the typhoon eye effect and ripple effect on people living with varying sizes of family, because people tend to live with larger families in Pakistani culture, and the larger family may either drain or provide buffering resource to mental issues. This study will also be one of the first of its kind to the mental health among adults in varying geographical locations in Pakistan. The findings of the research can help to pinpoints useful predictors that will help to provide targeted mental health support in vulnerable groups during the COVID-19 pandemic that still goes on in Pakistan.

## METHODS

### Study Context

The first case of COVID-19 in Pakistan was reported on February 26, 2020 in Karachi [23]. The largest city of Pakistan and the capital of Sindh, with a population of 16 million [24] has the high burden of disease as compared to other cities [1]. At the time of the study, Feb 26 to May 11 2020, there were 9480 cases in Karachi, representing 41.5% of the 22,820 total active cases in the entire country [23]. Hence Karachi was a clear epicenter in Pakistan at the time of the study.

### Data collection and sample

About 2.5 months into the outbreak, on May 4^th^ – 11^th^ 2020, we conducted an online survey of adults from all over Pakistan. On May 4^th^, 2020, when the survey started, the total number of confirmed cases of COVID-19 reached 21,501, and the death toll reached to 486 in Pakistan [23].

The study was approved by the Institutional Bioethical Committee of the University of Karachi (IBC KU -143/2020). The participants, after their consent, filled the online survey voluntarily. The survey promised the participants the confidentiality and anonymity of their responses. The participants could answer the survey in Urdu (the back-translated version) or English (the version developed originally).

### Variables

The participants reported their demographic characteristics such as age, gender, education, and marital status. They also reported their family size and daily exercise hours in the past week. We computed the distance from their geographical locations to Karachi, the COVID-19 epicenter of Pakistan.

The outcome variables included distress, anxiety, and, mental health. Distress was measured by K6, the six-item Kessler mental distress scale (0 = never, 4 = almost all of the time, α = 0.83) with the cut-off point of 13 [25]. Anxiety was measured by the Generalized Anxiety Disorder-7 scale (GAD-7) (0 = never, rarely, 3 = always; α = 0.88) with the cut-off point of 10 [26]. Mental health was assessed by Short Form-12 (SF-12) [27-29]. SF12 cover 8 sub-scales including physical functioning, physical role, body pain, general health, functionality, social functioning, emotional role, and mental health.

### Data analysis approach

We used Stata 16.0 to summarize the variables and to predict distress and anxiety by logistic regression and to predict mental health by ordinary least squares regression with a 95% confidence level.

## RESULTS

### Descriptive findings

The results showed that 47.6% of the 601 working adults were female, and 62.4% were younger than 29 years old, 26.0% were between 30 to 39 years, and 11.6% were 40 years or older. 67.7% of participants were single, 30.8% married, and 1.5% divorced. Most of the participants (70.5) had an undergraduate degree or higher with few participants (29.0) had a high school diploma (intermediate). On average, they exercised 0.77 hours each day with an SD of 0.79 hours. Overall, they had a family size of 6 with SD of 3 and resided 0.27 thousand kilometers away from Karachi, Sindh with SD of 0.51 thousand kilometers (see **Table 1**).

**Table 1.**
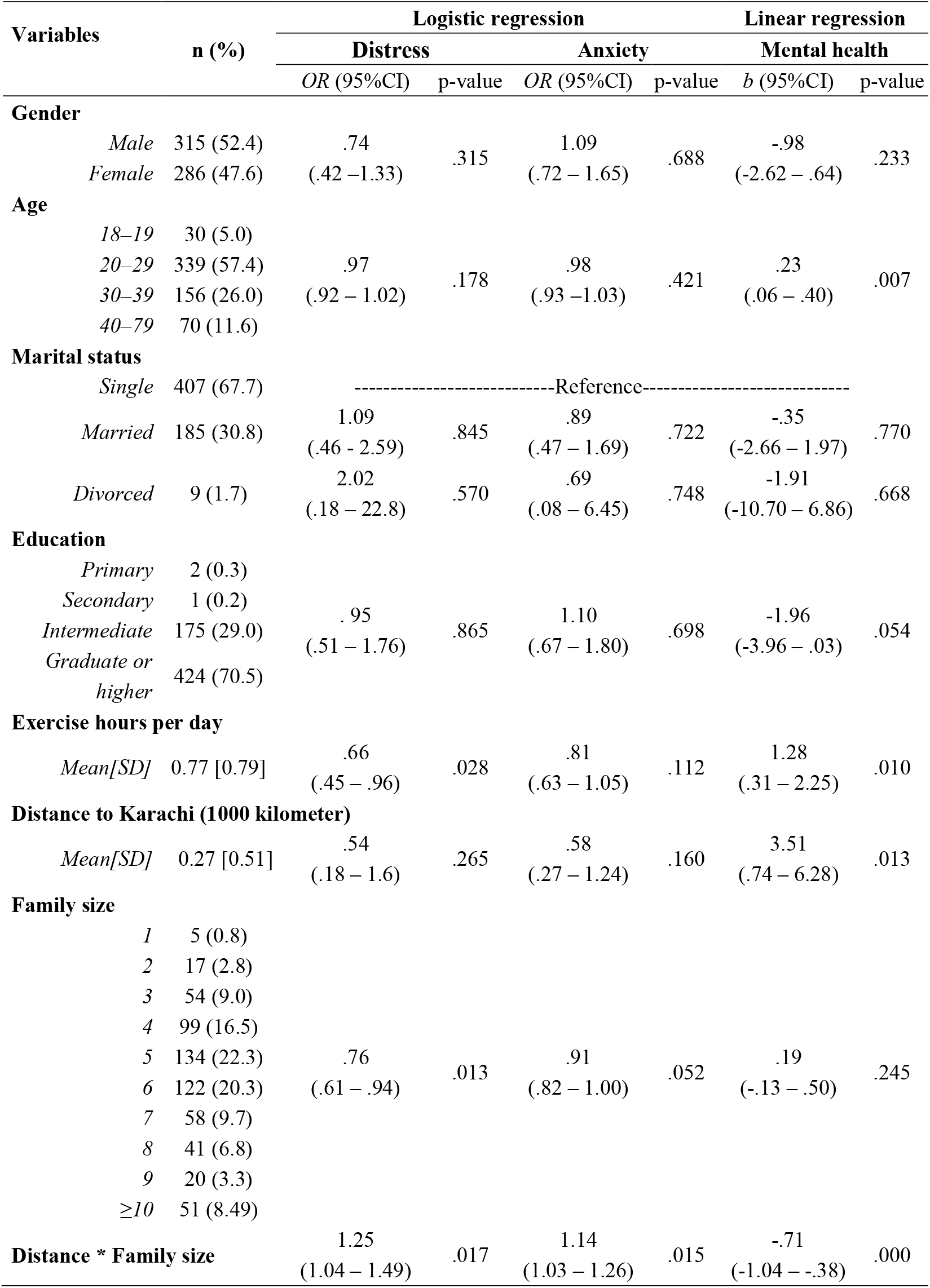
**Predicting working adults’ depression, anxiety, and mental health (N=601)**

| Variables |  | n (%) | Logistic regression |  |  |  | Linear regression |  |
| --- | --- | --- | --- | --- | --- | --- | --- | --- |
|  |  |  | Distress |  | Anxiety |  | Mental health |  |
|  |  |  | OR (95%CI) | p-value | OR (95%CI) | p-value | b (95%CI) | p-value |
| Gender |  |  |  |  |  |  |  |  |
|  | Male | 315 (52.4) | .74 | .315 | 1.09 | .688 | -.98 | .233 |
|  | Female | 286 (47.6) | (.42 – 1.33) |  | (.72 – 1.65) |  | (-2.62 – .64) |  |
| Age |  |  |  |  |  |  |  |  |
|  | 18–19 | 30 (5.0) |  | .178 |  | .421 |  | .007 |
|  | 20–29 | 339 (57.4) | .97 |  | .98 |  | .23 |  |
|  | 30–39 | 156 (26.0) | (.92 – 1.02) |  | (.93 – 1.03) |  | (.06 – .40) |  |
|  | 40–79 | 70 (11.6) |  |  |  |  |  |  |
| Marital status |  |  |  |  |  |  |  |  |
|  | Single | 407 (67.7) | -----Reference----- |  |  |  |  |  |
|  | Married | 185 (30.8) | 1.09 | .845 | .89 | .722 | -.35 | .770 |
|  |  |  | (.46 - 2.59) |  | (.47 – 1.69) |  | (-2.66 – 1.97) |  |
|  | Divorced | 9 (1.7) | 2.02 | .570 | .69 | .748 | -1.91 | .668 |
|  |  |  | (.18 – 22.8) |  | (.08 – 6.45) |  | (-10.70 – 6.86) |  |
| Education |  |  |  |  |  |  |  |  |
|  | Primary | 2 (0.3) |  | .865 |  | .698 |  | .054 |
|  | Secondary | 1 (0.2) |  |  |  |  |  |  |
|  | Intermediate | 175 (29.0) | .95 |  | 1.10 |  | -1.96 |  |
|  | Graduate or | 424 (70.5) | (.51 – 1.76) |  | (.67 – 1.80) |  | (-3.96 – .03) |  |
|  | higher |  |  |  |  |  |  |  |
| Exercise hours per day |  |  |  |  |  |  |  |  |
|  | Mean[SD] | 0.77 [0.79] | .66 | .028 | .81 | .112 | 1.28 | .010 |
|  |  |  | (.45 – .96) |  | (.63 – 1.05) |  | (.31 – 2.25) |  |
| Distance to Karachi (1000 kilometer) |  |  |  |  |  |  |  |  |
|  | Mean[SD] | 0.27 [0.51] | .54 | .265 | .58 | .160 | 3.51 | .013 |
|  |  |  | (.18 – 1.6) |  | (.27 – 1.24) |  | (.74 – 6.28) |  |
| Family size |  |  |  |  |  |  |  |  |
|  | 1 | 5 (0.8) |  | .013 |  | .052 |  | .245 |
|  | 2 | 17 (2.8) |  |  |  |  |  |  |
|  | 3 | 54 (9.0) |  |  |  |  |  |  |
|  | 4 | 99 (16.5) |  |  |  |  |  |  |
|  | 5 | 134 (22.3) | .76 |  | .91 |  | .19 |  |
|  | 6 | 122 (20.3) | (.61 – .94) |  | (.82 – 1.00) |  | (-.13 – .50) |  |
|  | 7 | 58 (9.7) |  |  |  |  |  |  |
|  | 8 | 41 (6.8) |  |  |  |  |  |  |
|  | 9 | 20 (3.3) |  |  |  |  |  |  |
|  | ≥10 | 51 (8.49) |  |  |  |  |  |  |
| Distance * Family size |  |  | 1.25 | .017 | 1.14 | .015 | -.71 | .000 |
|  |  |  | (1.04 – 1.49) |  | (1.03 – 1.26) |  | (-1.04 – -.38) |  |

### Descriptive and comparative findings on the outcome variables

About one-tenth of participants surpassed the cut-off of distress (9.2%) and about one-fifth of participants surpassed that of anxiety (19.0%). By comparing our findings with those in 11 studies using similar measurements, we found that overall the mental health conditions of Pakistani adults were comparable or less than those in several samples in China, Spain, and Italy with one exception (see **Table 2** for a summary). Anxiety disorder in our sample was higher than that in a sample of adults in China in late Feb 2020 [30].

**Table 2.** **The comparisons of adults’ distress and anxiety issues during the COVID-19 pandemic across studies**

| Measure | Sample description; data collection time | Prevalence | Comparison with this study | Source |
| --- | --- | --- | --- | --- |
| **Distress** | **This study** | **9.2%** | **-** |  |
| Kessler-6 | 369 adults in China, Feb 20–21, 2020 | 6.2% | -3.0% (-6.3% to 0.6%)<br>$\chi^2(1)=2.8$, $p=0.10$ | [4] |
| Kessler-10 | 500 adults in Italy, April 10–13, 2020 | 18.6% | 9.4% (5.5% to 13.3%)<br>$\chi^2(1)=22.2$, $p<0.0001$ | [6] |
| Kessler-6 | 1599 adults in China, Feb 1–4, 2020 | Mean (SD): 7.7<br>($\pm 7.7$) | 2.2% (1.49% - 2.8%)<br>T (2198) = 6.4, $p<0.0001$ | [46] |
| Kessler-6 | 2032 adults in the U.S., late April 2020 | 27.7% | 18.5% (15.3% to 21.4%)<br>$\chi^2(1)=88.3$, $p<0.0001$ | [35] |
| **Anxiety** | **This study** | **19.0%** | **-** |  |
| GAD-2 | 3088 adults in 32 provinces of China, Feb 20–27, 2020 | 13.2% | -5.83% (-2.6% to -9.3%)<br>$\chi^2(1)=13.9$, $p=0.0002$ | [30] |
| GAD-2 | 3480 adults in Spain, March 21–27, 2020 | 21.6% | 2.3% (-1.3% to 5.5%)<br>$\chi^2(1)=1.6$, $p=0.21$ | [36] |
| GAD-7 | 103 adults in China, Feb 10–28, 2020 | 22.3% | 3.3% (-4.4% to 12.7%)<br>$\chi^2(1)=0.6$, $p=0.44$ | [47] |
| GAD-7 | 98 adults in Zhongshan, Guangdong in China, Feb 15–29, 2020 | 23.4% | 4.4% (-3.6% to 14.1%)<br>$\chi^2(1)=1.03$, $p=0.31$ | [34] |
| GAD-7 | 4872 adults in China, Jan 31–Feb 2, 2020 | 22.6% | 3.6% (.1% - 6.8%)<br>$\chi^2(1)=4.0$, $p = .045$ | [48] |
| GAD-2 | 1577 adults in Wuhan, China, Feb 18–24, 2020 | 23.8% | 4.8% (.9% - 8.5%)<br>$\chi^2(1)=5.7$, $p = .017$ | [49] |
| GAD-7 | 1556 seniors older than 60 years in China | 37.1% | 18.1% (14.0% - 21.9%)<br>$\chi^2(1)=65.2$, $p < .0001$ | [50] |

### Predictors of distress, anxiety, and mental health

The distance to the epicenter of COVID-19 in Pakistan negatively predicted the mental health of adults, but the relationship depended on their family size (b=-0.71; 95% CI: -1.04 to -0.38; P=0.000). Margin analysis showed that the distance to the epicenter positively predicted mental health for adults in a small family (e.g. at a single family: b=2.79; 95% CI: 0.28 to 5.30; P=0.039). In contrast, the distance to the epicenter negatively predicted mental health for adults in large family (e.g. at an 8-member family: b=-2.19; 95% CI: -3.85 to -0.54; P=0.009). Similarly, the relationship of the distance to the epicenter and mental health disorders of adults also depended on their family size (OR=1.25; 95% CI: 1.04 to 1.49; P=0.017 for distress, and OR=1.14; 95% CI: 1.03 to 1.26; P=0.015 for anxiety). Margin analysis showed that the distance to the epicenter positively predicted distress disorder for adults in large families (e.g. at an 8-member family: OR=0.065; 95% CI: 0.032 to 0.098; P=0.000) and anxiety disorder (e.g. at an 8-member family: OR=0.066; 95% CI: 0.008 to 0.12; P=0.026) (see **Figure 1**).

**Figure 1.**
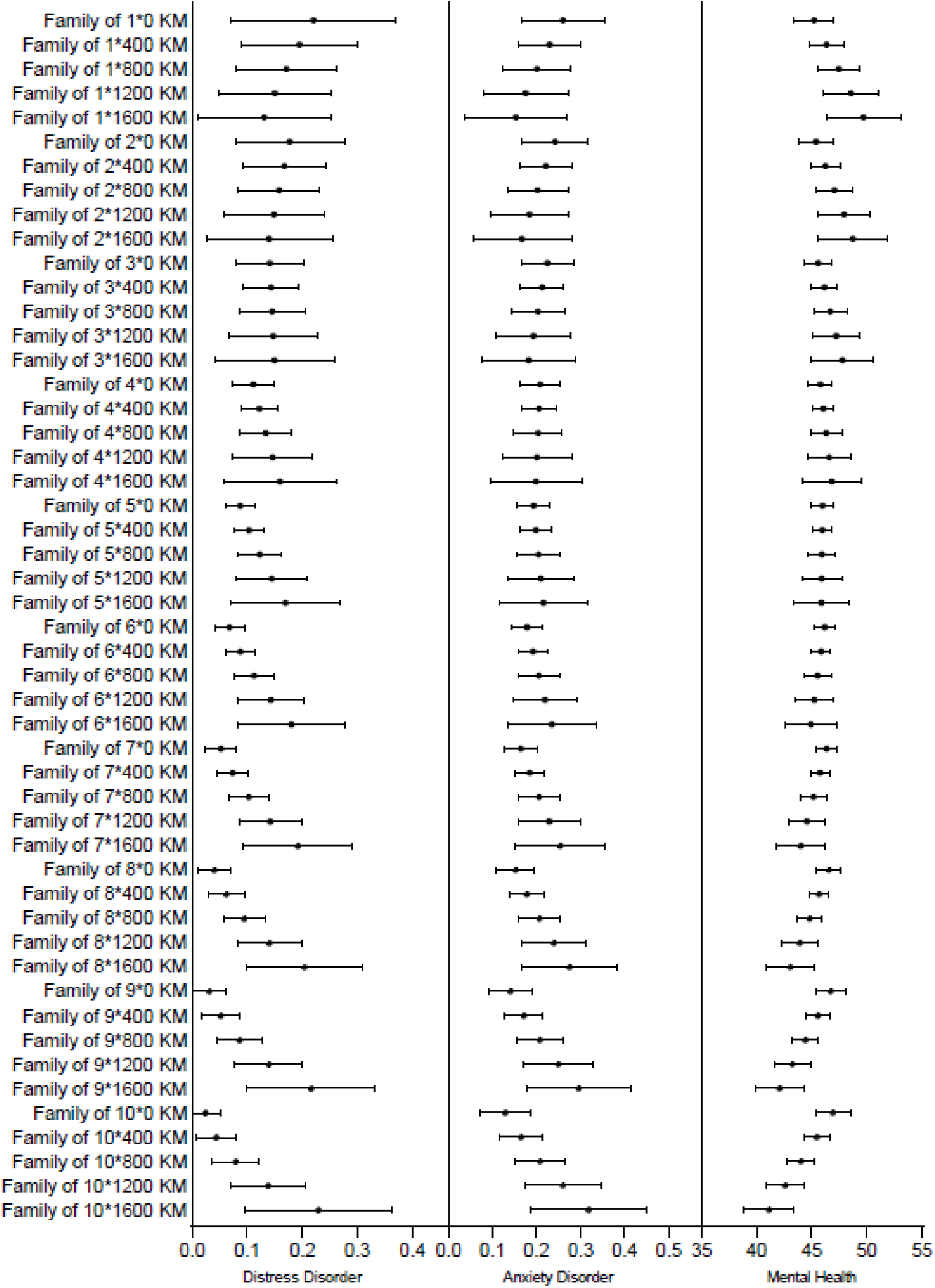
Predicted value and 95% confidence intervals of distress, anxiety and mental health by family size and distance to the epicenter

In addition, adults who exercised more have better mental health (b=1.28; 95% CI: 0.31 to 2.25; P=0.010) and less likely to experience distress disorders (OR=0.66; 95% CI: 0.45 to 0.96; P=0.028). The results also suggest that older the person, the improved mental health (b=0.28; 95% CI: 0.06 to 0.40; P=0.007).

## DISCUSSION

Pandemics have a myriad impact on the mental health of people [31]. In the recent outbreak of COVID-19, it has been reported that COVID-19 itself with many other factors has increased mental issues in various countries [4, 8, 32-34]. To the best of our knowledge this is the first study to examine the psychological typhoon eye effect or ripple effect from the epicenter among Pakistani adults. The findings of psychological distress and anxiety scales revealed the prevalence of moderate distress and anxiety in our sample. Compared to other recently published studies, the results showed that the rate of anxiety and distress among Pakistani adults was greater as compared to those in China [30], whereas lower as compared to Italy, Spain and United States [6, 35, 36]. These differences might be due to less number of reported cases and deaths as compared to the countries that have high distress and anxiety. With regard to the variables associated with distress, anxiety and mental health in Pakistan, family size and exercise were worth noting predictors found in our sample. Previous literature revealed that geographical distance from the epicenter as an important prognosticator during catastrophic events [19]. In the present study the findings overall showed that participants residing distantly from the epicenter have better mental health with less distress and anxiety, supporting the ripple effect rather than the typhoon eye effect [7, 17]. However, the association can diverge based on individuals’ family size. The mental disorder decreased by the distance to the epicenter for individuals in small families, indicating the typhoon eye effect. By contrast, mental disorder increased by the distance to the epicenter for individuals in larger families, showing the ripple effect.

Our results on the ripple effect vs. typhoon eye effect, together with other studies on the same topic in China, Brazil, and Peru [7, 17, 20], suggest the prediction of these two opposing theories may differ based on the countries studied. Such differences are understandable, as countries vary in their geography, media and social media reporting, medical systems, cultures, the availability of personal protective equipment (PPE), labor and employment conditions, the policies of lockdown, the ease of working from home, and maintaining a living in a pandemic, and the information in both mainstream and social media, etc. [5]. The results therefore suggest we need to identify the predictive effect of the ripple effect vs. typhoon eye effect as a predictive model of mental health in individual countries during the Covid-19 pandemic. Our results suggest such studies can focus on identifying relevant contingency factors in individual countries. In Pakistan, we focused on family size, and the possible explanation for family size to reverse the effect of the ripple effect vs. typhoon eye effect might be due to socio-economic burden on the families due to lockdown in the country. As studies have shown that financial constraints or economic hardships not only increased behavioral problems but also damaged the physical and mental health status of individuals and their families [37]. Thus, our findings identify family size as a critical contingency factor in the prediction of typhoon eye effect and ripple effect. Future research can focus on identifying unique contingency factors in the country of the study.

As in previous studies in Brazil, China and Iran [4, 5, 20], our sample also identified exercise hours as one of the predictors of distress, anxiety and mental health during COVID-19. The results showed that participants who put in more hours for physical activity in terms of daily exercise in their routine, had better mental health and were less likely to develop distress and anxiety symptoms. Many studies have reported that performing daily exercise might have an impact on anxiety and distress symptoms [38, 39]. Due to the sedentary life style in COVID-19 pandemic, it has been observed that people gave less attention to their physical health than in normal circumstances [40]. Thus, particularly in this pandemic era when people were extra stressed, adding physical activity in the daily routine can play a role in reducing distress and anxiety. In comparisons with the recent studies of Iran, China and Brazil [41-43], age also predicted mental health in the Pakistani population. The results showed that older people had better mental health, which might be due to the joint family system in Pakistan. It has been reported that traditional family systems, such as those in South Asia, can contribute to healthier mental states among older people as compared to those living in smaller nuclear family systems [44]. The positive attitude from a lack of information about COVID-19 could also be another factor for better mental health of older people [45]. As compared to older people, younger people rely more on social media and the internet that have helped to spread negative information on the pandemic [18, 30].

The overall findings of the present study can help to identify the vulnerable individuals in Pakistan during this crisis of COVID-19. Exercise, family size, age and distance to the epicenter are the key predictors of distress, anxiety and mental health in Pakistan during this pandemic. More specifically the relationship of the geographical distance to the epicenter with distress, anxiety and mental health represented the ripple effect in large families. However, the relationship varied depending on family size and showed the typhoon eye effect in small families. Thus, results suggest psychiatrists and mental health care providers that the geographical distance to epicenter, with an important contingency of facility size, can play a major role in screening of people with high risk.

This study had some limitations. During the survey dates, the total active cases of COVID-19 in Pakistan had yet to reach its peak, and the situation continues to evolve. In addition, as the study was conducted through an online questionnaire, selective participation and coverage area error might be present. While we aimed for broad coverage of the adults in various parts of Pakistan, we do not claim our sample to be representative of the adults’ population in Pakistan.

## CONCLUSION

In conclusion the present study uncovered the prevalence of distress and anxiety disorders in Pakistani adults during COVID-19. The results indicate that geographical distance is a crucial factor in screening vulnerable groups and suggest future studies to examine the use of the typhoon eye effect or ripple effect in terms of identifying mentally vulnerable people with a focus to identify the relevant contingency factors.

## Data Availability

The datasets presented in this article are not available. Requests to access the datasets should be directed to corresponding author.

## DECLARATIONS

### Ethics Approval and Consent to Participate

The study was approved by Institutional Bioethical Committee, University of Karachi (approval no. IBC KU-143/2020). Each study participant filled the online questionnaire voluntarily after their consent.

### Consent for Publication

Not applicable

### Competing Interest

The authors declare that there are no potential conflicts of interest with respect to the research, authorship, and/or publication of this article.

### Funding

None

### Authors Contribution

**T. L**.: Investigation (data collection), Writing – Original, Writing – Review & Editing

**J. C**.: Investigation, Formal analysis, Investigation, Writing – Original, Writing – Review & Editing, Validation

**M. T**.: Investigation (data collection), Resources, Writing – Review & Editing

**T. A. L**.: Investigation (data collection), Writing – Review & Editing

**B. C**.: Visualization; Writing – Review & Editing

**J. L**.:

**S. X. Z**.: Conceptualization, Investigation, Methodology, Formal analysis, Writing – Original, Writing – Review & Editing, Supervision

All authors read and approved the final manuscript.

## Acknowledgements

We thank the Institutional Bioethical Committee, the University of Karachi for support. We also thank all individuals who have devoted their time to participate in this study.

## Notes

### Competing Interest Statement

The authors have declared no competing interest.

### Clinical Trial

Not applicable

### Author Declarations

Institutional Bioethical Committee University of Karachi Approval No. IBC KU-143/2020

